# Association between blood pressure and new onset of chronic kidney disease in non-diabetic Japanese adults: a population-based longitudinal study from 1998 to 2023

**DOI:** 10.1101/2023.12.06.23299067

**Authors:** Yukari Okawa, Toshiharu Mitsuhashi

**Affiliations:** Department of Public Health and Welfare, Zentsuji City Hall, Zentsuji-city, Kagawa, Japan; Center for Innovative Clinical Medicine, Okayama University Hospital, Okayama, Japan

**Author notes:** **Corresponding author:** Yukari Okawa, Department of Public Health and Welfare, Zentsuji City Hall, 2-1-1 Bunkyo-cho, Zentsuji City, Kagawa 765-8503, Japan.

**Keywords:** Hypertension, Chronic Kidney Diseases, Longitudinal Studies, Risk Factors

## Abstract

**Background and Aims:** Hypertension is a risk factor for developing chronic kidney disease (CKD). Studies of adult participants in the USA reported that hypertension increased the risk of developing CKD even in the non-diabetic population. However, studies in non-diabetic populations are limited and additional studies in other races are required. This study aimed to examine the relationship between hypertension and the development of CKD in non-diabetic Asian adults.

**Methods and Results:** In this longitudinal study, non-diabetic Japanese adults who took annual checkups from 1998 to 2023 were included. CKD was defined as <60 mL/min/1.73 m^2^, and hypertension was classified into four levels according to the guidelines of the American College of Cardiology/American Heart Association. The Weibull accelerated failure time model was selected because the proportional hazards assumption was violated. Of the 7 363 (men: 40.3%) people in the final cohort, 2 498 (men: 40.1%) developed CKD after a mean follow-up of 7.99 years. Elevated blood pressure (BP) and hypertension stage 2 had a 9% (95% confidence interval [CI]: 1%–16%) and 11% (95% CI: 5%–17%) shorter survival time to CKD onset, respectively, than normal BP. Hypertension stage 1 also had a shorter survival to CKD onset by point estimate, but all 95% CIs crossed 1 in all models.

**Conclusions:** In a relatively healthy Asian population without diabetes, controlling BP to an appropriate range reduces the risk of developing CKD.

## 1 Introduction

Chronic kidney disease (CKD) is a well-known public health problem that affects 9.1% of the global population[1]. CKD is defined as an abnormality in renal structure or function that affects health for longer than 3 months[2]. CKD is sometimes referred to as a silent killer because it progresses without symptoms to an irreversible state. Once CKD progresses to end-stage renal disease, treatment options are limited to expensive dialysis and kidney transplantation, with limited access to treatment[3]. Therefore, early detection and prevention of CKD are critical.

To date, a variety of risk factors for CKD have been reported, including diabetes, a high body mass index (BMI), hypertension, non-alcoholic fatty liver disease, hepatitis B and C virus infection, a Western-type dietary pattern, and air pollution[4–9]. Among those, diabetes is the leading cause of developing CKD, accounting for 30.7% of CKD disability-adjusted life-years[1,4]. However, risk factors for the development of CKD in a non-diabetic population have not yet been fully investigated.

According to a meta-analysis that integrated the results of four cohort studies, men with hypertension had a 2.14-fold higher risk (95% confidence interval [CI]: 1.68–2.72) and women with hypertension had a 1.54-fold higher risk (95% CI: 1.37–1.74) of developing CKD than those with normal blood pressure (BP)[5]. However, the four cohort studies used multiple definitions of CKD and hypertension and included individuals with and without diabetes, which may have affected the results[10–12]. Therefore, the relationship between BP levels and the subsequent development of CKD in a non-diabetic population needs to be investigated in more detail.

The Atherosclerosis Risk in Communities study, which followed 10 096 middle-aged non-diabetic American adults for 9 years, reported that after adjustment for age, sex, and race, participants with hypertension were 1.99 times (95% CI: 1.69–2.35) more likely to develop CKD than those with normal BP[13]. In this previous study, hypertension was dichotomized into prevalent (systolic blood pressure [SBP]/diastolic blood pressure [DBP] ≥130/≥85 mmHg or the use of BP medications) and non-prevalent (SBP/DBP <130/<85 mmHg) categories[13]. Furthermore, racial disparities in the prevalence of hypertension have been reported, with a higher prevalence in blacks and other races (other than whites, blacks, Hispanics, and Asians) than in whites[14,15]. Therefore, to understand the effect of BP on the development of CKD, examining the relationship between more detailed BP categories and the development of CKD in a single racial population is important. The American College of Cardiology/American Heart Association (ACC/AHA) recommends classifying BP in adults into four levels, which allow more precise categorization than dichotomized BP classification. These levels are as follows: normal (SBP <120 and DBP <80 mmHg), elevated (SBP of 120–129 and DBP <80 mmHg), hypertension stage 1 (SBP of 130–139 or DBP 80–89 mmHg), and hypertension stage 2 (SBP ≥140 or DBP ≥90 mmHg)[15]. These four levels of ACC/AHA BP classification can be used to study the effect of BP on the development of CKD in more detail.

Therefore, this study aimed to examine the relationship between the ACC/AHA BP classification and new onset of CKD in a non-diabetic Japanese middle-aged and older population.

## 2 Methods

### 2.1 Study population and data source

This ongoing, open cohort study used administrative annual health checkup data from Zentsuji City from 6 April 1998 to 19 April 2023. This city is located around the center of Kagawa Prefecture in Japan, with a population of 30 431 (men: 49.7%, aged ≥60 years: 38.1%) as of 1 April 2023[16]. All data were extracted from the Zentsuji City database on 6 July 2023. The database used was the same as that used in our previous study[17].

The checkup was conducted annually for citizens ≥40 years by fiscal year age according to the protocol established by the Ministry of Health, Labour and Welfare[18]. In addition, to promote the health of young people, the city expanded the age range for checkup to 35–39 years by fiscal year age on a trial basis only in the fiscal years 1998 and 1999. Each year, 30%–40% of the eligible population receives a checkup. The checkup includes anthropometric measurements, a BP test, a blood test, a urinalysis, and a self-reported questionnaire asking about participants’ lifestyles, such as drinking and smoking status. The total number of participants of this checkup who were included in this study was 15 501 (men: 40.7%).

On the basis of the study aim to investigate the relationship between hypertension and the subsequent development of CKD among non-diabetic Japanese adults, the following participants were excluded from the analysis: non-Japanese and those with missing renal information, missing glycated hemoglobin (HbA1c) values, missing SBP or DBP values, prevalent CKD at study entry, missing information on glycosuria at study entry or prevalent glycosuria ≥1+ at study entry, prevalent diabetes at study entry, or only a single observation. Participants who developed diabetes during the follow-up period were treated as censored. An HbA1c value of ≥6.5% was treated as diabetes according to the American Diabetes Association criteria[19].

### 2.2 Variables

#### 2.2.1 Outcome

In the analysis, the onset of CKD was the outcome variable. The estimated glomerular filtration rate (eGFR) was used as a measure of renal function. To mitigate the effect due to race, we used the three-variable Japanese equation to calculate the eGFR as follows : *eGFR* (*mL*/*min*/1.73*m*^2^) = 194 × *serum creatinine* (*mg*/*dL*)^-1.094^ × *age* (*years*)^-0.287^ (× 0.739 *if female*)[20–22]. Prevalent CKD was defined as an eGFR <60 mL/min/1.73 m^2^ according to the Kidney Disease Improving Global Outcome. Serum creatinine concentrations (mg/dL) were measured to two decimal places using enzymatic methods.

#### 2.2.2 Exposure

The exposure variable was prevalent hypertension. SBP and DBP were used to define normal BP and hypertension by following the ACC/AHA guidelines: normal (SBP <120 and DBP <80 mmHg, reference), elevated (SBP of 120–129 and DBP <80 mmHg), hypertension stage 1 (SBP of 130–139 or DBP of 80– 89 mmHg), and hypertension stage 2 (SBP ≥140 or DBP ≥90 mmHg)[15].

#### 2.2.3 Other covariates

To reduce potential bias, sociodemographic and modifiable lifestyle factors were adjusted for. Sociodemographic factors included age and the residential district. Age was coded into three groups of 34–59, 60–69, and 70–100 years. The residential districts of the city were as follows: East, West, Central, South, Fudeoka, Tatsukawa, Yogita, and Yoshiwara.

Modifiable lifestyle factors were the self-reported drinking status, the self-reported smoking status, overweight or obesity, dyslipidemia, HbA1c values, proteinuria, and glycosuria. The self-reported drinking status was dichotomized into “non- or seldom-drinker” and “drinker.” The self-reported smoking status was grouped into “non- or ex-smoker” and “smoker”. BMI was calculated as weight (kg) divided by height (m) squared, and classified into “normal weight (BMI <25 kg/m^2^)” and “overweight or obese (BMI ≥25 kg/m^2^)”[23]. The participants were classified as dyslipidemic if they met any of the following conditions: serum low-density lipoprotein cholesterol concentrations ≥140 mg/dL, serum high-density lipoprotein cholesterol concentrations <40 mg/dL, or serum triglyceride concentrations ≥150 mg/dL[24]. HbA1c was standardized to National Glycohemoglobin Standardization Program (NGSP) values (%) by the officially certified equation because HbA1c was reported in Japan Diabetes Society (JDS) units (%) from 1998 to 2012: *HbA1c_NGSP_* (%) = 1.02 × *HbA1c_JDS_*(%) + 0.25[25,26]. Proteinuria and glycosuria were dichotomized into “non-prevalent (none or ±)” and “prevalent (≥1+)”.

The questionnaire was revised by the Ministry of Health, Labour and Welfare in the middle of the fiscal year 2012, and questions regarding antihypertensive medication use, a history of heart disease (e.g., angina pectoris and myocardial infarction), and a history of stroke (e.g., cerebral hemorrhage and cerebral infarction) were added to the questionnaire. All types of antihypertensive medications were included but generic names were not asked. The majority of observations on the use of antihypertensive medication (missing 58.9% of the total observations), a history of heart disease (missing 60.4% of the total observations), and a history of stroke (missing 60.4% of the total observations) were missing. Therefore, these variables could not be included in the current analysis. In the fiscal year 2013, 42.4% of questionnaire respondents reported the use of antihypertensive medications, 8.86% reported a history of heart disease, and 5.15% reported a history of stroke.

### 2.3 Statistical analysis

The participants’ characteristics are summarized by BP classifications. Continuous variables are expressed as the mean and standard deviation (SD). The person-years at risk were calculated from the date of the first observation to the occurrence of an event or the development of diabetes, or to the end of the last observation during the follow-up if no event or diabetes occurred. The participants’ characteristics during the follow-up are expressed as the number who developed CKD, total person-years, and incidence rate per 1 000 person-years.

The proportions of missing values of variables were 0.02% for overweight or obesity, 29.6% for the self-reported drinking status, 23.9% for the self-reported smoking status, 21.3% for dyslipidemia, and 1.38% for the residential district. The imputed results were selected in this analysis because missing values were assumed to be missing at random. All missing values were complemented by multiple imputation using chained equations with 40 imputations[27,28]. Binary variables (overweight or obesity, self-reported drinking status, self-reported smoking status, and dyslipidemia) were imputed using logistic regression and categorical variables (residential district) by multinominal logistic regression.

A Kaplan–Meier curve was created. The log–log plots and Schoenfeld residual results showed that the proportional hazards (PH) assumption was violated[29]. The Weibull accelerated failure time model was selected according to the Akaike and Bayesian information criteria because the PH assumption was violated, and all variables, except for sex, varied by time[30,31]. The time ratio and 95% CI were indicators of an association between exposure and outcome. A time ratio <1 indicates a shorter survival time to CKD onset than the reference group. An example of this ratio is as follows: a time ratio of 0.8 indicates that participants with hypertension have a 0.8 times longer survival than those with normal BP. To avoid the Table 2 fallacy, only estimates of the primary exposure were presented[32].

Covariates were selected on the basis of previous studies[5]. Model 1 was adjusted for sex and age group. Model 2 was further adjusted for overweight or obesity, the self-reported drinking status, and the self-reported smoking status. Model 3 was further adjusted for dyslipidemia and HbA1c values. Model 4 was further adjusted for the residential district. A multiplicative term was added to Models 1–4 for stabilizing models when there was an interactive relationship between the BP category and the following variables: sex, age category, overweight or obesity, the self-reported drinking status, the self-reported smoking status, dyslipidemia, and HbA1c values.

A two-tailed p value <0.05 was considered statistically significant. STATA/SE 18.0 (StataCorp, College Station, TX, USA) was used to conduct all statistical analyses and to create a Kaplan–Meier curve. The geographic data were downloaded from the website of the National Statistics Center, Japan[33]. A flow chart of the participants and the district map of Zentsuji City were created in Python 3.11.5[34–36]. This study followed the Strengthening the Reporting of Observational Studies in Epidemiology reporting guidelines[37].

### 2.4 Sensitivity analyses

We conducted two sensitivity analyses. The first sensitivity analysis used SBP and DBP as exposure variables. SBP was categorized into the following six groups: <100, 100–119 (reference), 120–129, 130– 139, 140–149, and ≥150 mmHg. DBP was classified into the following five groups: <70, 70–79 (reference), 80–89, 90–99, and ≥100 mmHg. To reduce potential reverse causation, the second sensitivity analysis excluded participants with proteinuria at study entry. All participants had information on proteinuria at study entry.

### 2.5 Ethics

All data were retrieved from the administrative database of Zentsuji City and anonymized prior to receipt. The Ethics Committee of Okayama University Graduate School of Medicine, Dentistry and Pharmaceutical Sciences and Okayama University Hospital approved this study and waived the need for informed consent (No. K1708-040). All study procedures were conducted in accordance with the Declaration of Helsinki and Japanese Ethical Guidelines for Medical and Biological Research Involving Human Subjects.

### 2.6 Conflicts of interest

This research did not receive any specific grant from funding agencies in the public, commercial, or not-for-profit sectors. YO declares a relationship with Zentsuji City that includes employment. TM declares no potential conflict of interest associated with this manuscript.

## 3 Results

### 3.1 Main analysis

Of 15 501 (men: 40.7%, mean age at study entry: 62.3 years [SD: 12.5]) initial participants, 7 363 (men: 40.3%, mean age at study entry: 60.3 years [SD: 11.5]) remained in the final cohort (Figure 1). After a 25-year follow-up, with a mean follow-up per participant of 7.99 years, 2 498 (men: 40.1%) participants developed CKD.

**Figure 1.**
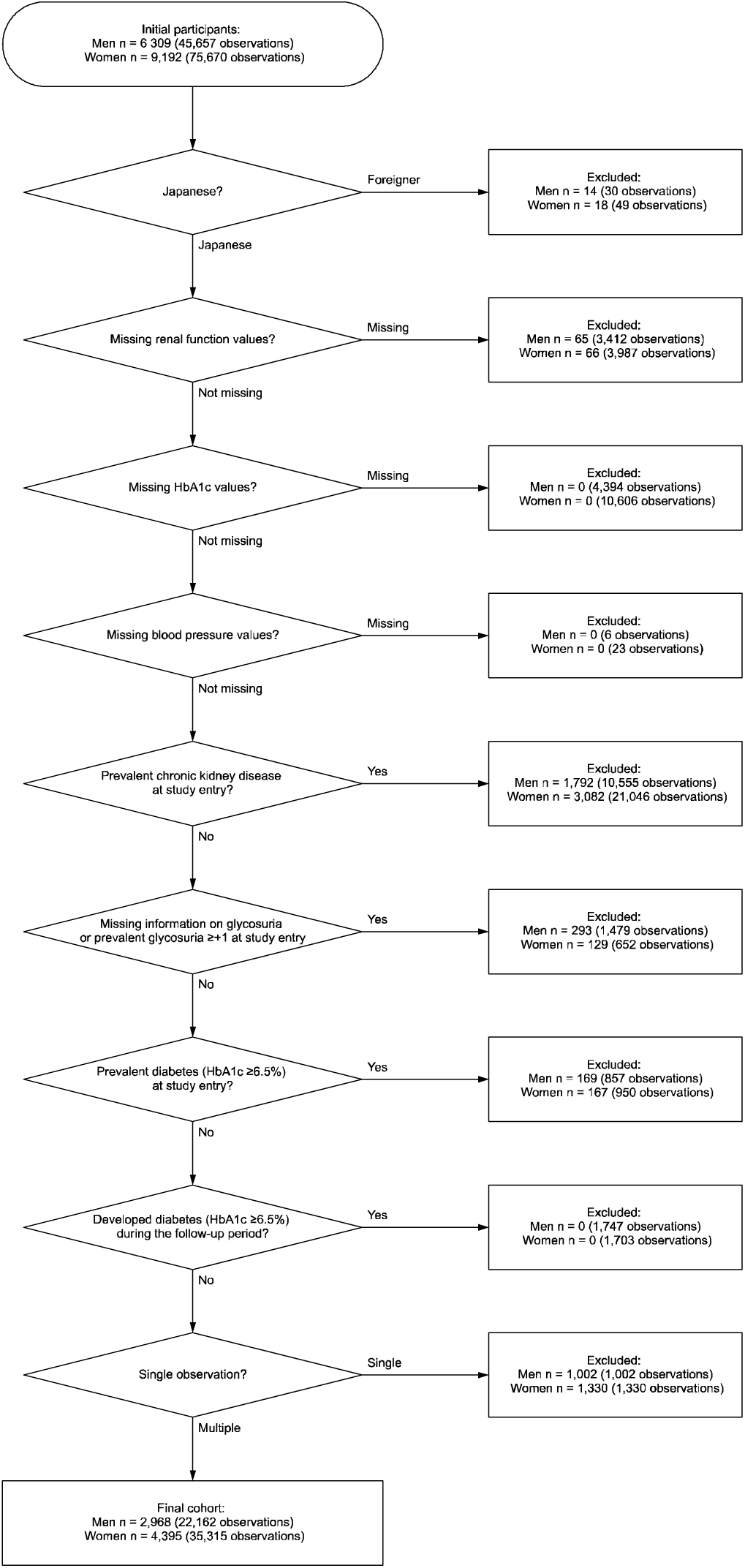
Participants’ flow chart of the study cohort.

Figure 2 shows the map of the Zentsuji City district. The Kaplan–Meier curves stratified by the ACC/AHA BP classification are presented in Figure 3. Table 1 shows the participants’ characteristics stratified by the ACC/AHA BP classification. Participants with higher BP categories were more likely to be men, older, overweight or obese, and/or dyslipidemic. Participants with elevated BP had the shortest follow-up of all BP categories (14.9% of the total).

**Figure 2.**
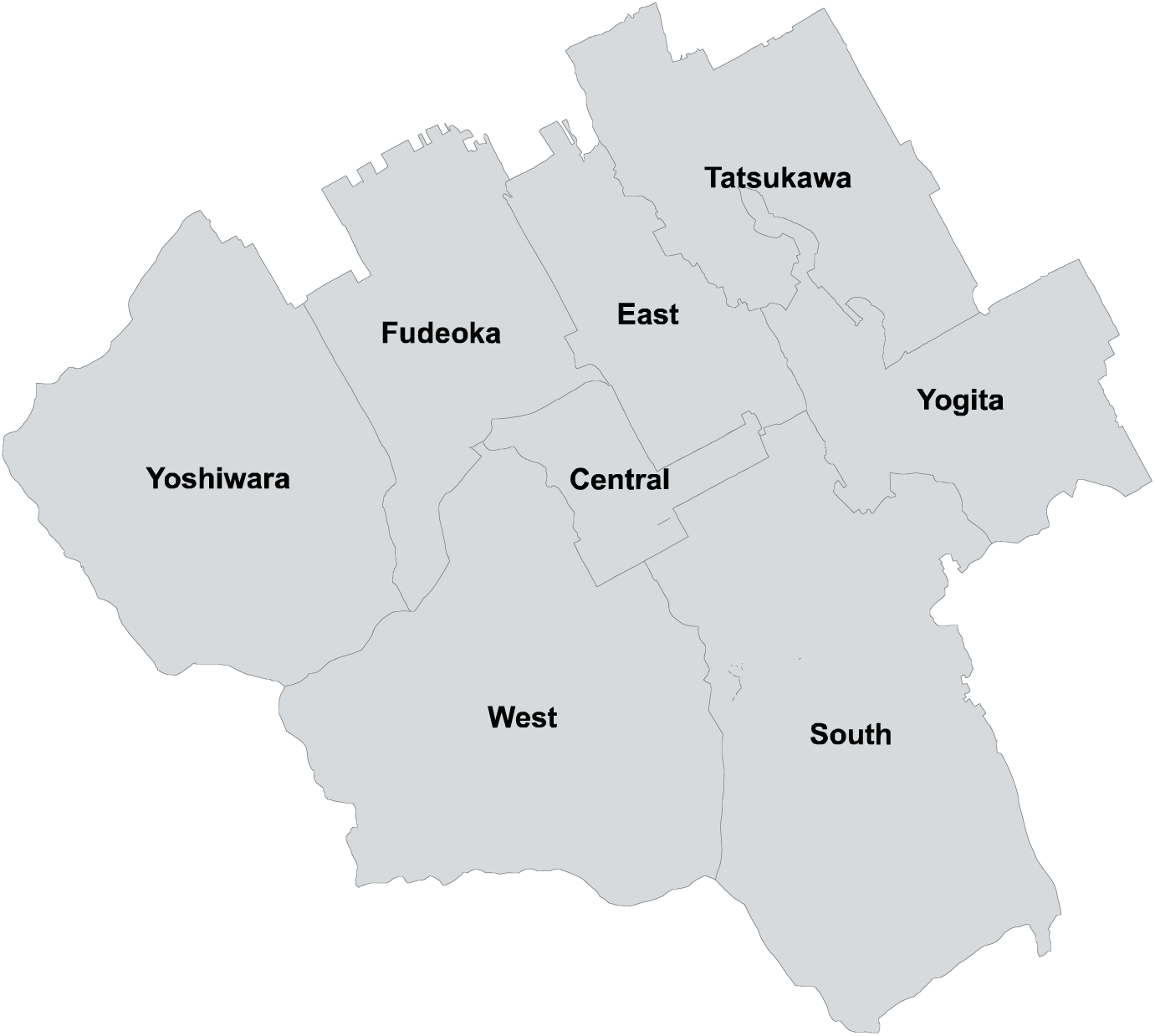
Map of Zentsuji City district.

**Figure 3.**
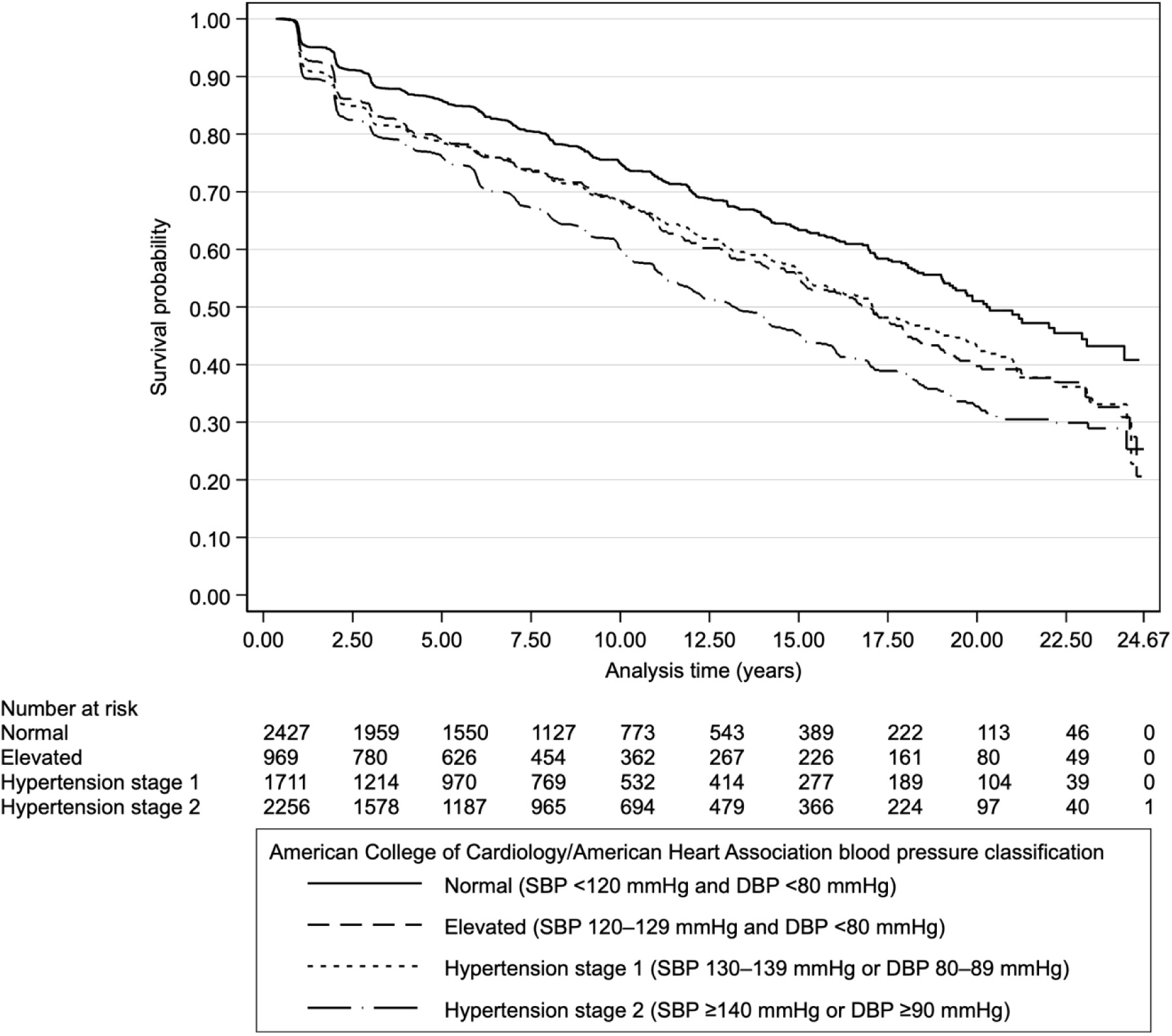
Kaplan–Meier survival estimates by the American College of Cardiology/American Heart Association blood pressure classification.

**Table 1.** Descriptive statistics of all observations of 7 363 non-diabetic Japanese citizens of Zentsuji city stratified by the ACC/AHA blood pressure classification (1998–2023).

|  | SBP (mmHg)<br>DBP (mmHg) | ACC/AHA blood pressure classification |  |  |  |  |  |  |  |  |  |  |  | Total |  |  |
| --- | --- | --- | --- | --- | --- | --- | --- | --- | --- | --- | --- | --- | --- | --- | --- | --- |
|  |  | Normal |  |  | Elevated |  |  | Hypertension stage 1 |  |  | Hypertension stage 2 |  |  |  |  |  |
|  |  | <120 and<br><80 |  |  | 120–129 and<br><80 |  |  | 130–139 or<br>80–89 |  |  | ≥140 or<br>≥90 |  |  |  |  |  |
|  |  | Failure | PY | IR* | Failure | PY | IR* | Failure | PY | IR* | Failure | PY | IR* | Failure | PY | IR* |
| Time at risk |  |  |  |  |  |  |  |  |  |  |  |  |  |  |  |  |
| Total |  |  | 19 834.1 |  |  | 8 787.8 |  |  | 13 393.1 |  |  | 16 841.6 |  |  | 58 856.5 |  |
| Minimum/Maximum |  |  | 0.34/24.4 |  |  | 0.39/24.5 |  |  | 0.29/23.0 |  |  | 0.37/21.2 |  |  | 0.34/24.7 |  |
| Mean |  |  | 5.14 |  |  | 2.84 |  |  | 3.45 |  |  | 4.46 |  |  | 7.99 |  |
| Median |  |  | 3.19 |  |  | 1.99 |  |  | 2.13 |  |  | 3.02 |  |  | 6.44 |  |
| Failure/IR* |  | 613 |  | 30.9 | 383 |  | 43.6 | 576 |  | 43.0 | 926 |  | 55.0 | 2 498 |  | 42.4 |
| Variables |  |  |  |  |  |  |  |  |  |  |  |  |  |  |  |  |
| Sex |  |  |  |  |  |  |  |  |  |  |  |  |  |  |  |  |
| Female |  | 377 | 13 186.1 | 28.6 | 233 | 5 629.7 | 41.4 | 340 | 7 993.0 | 42.5 | 547 | 9 813.4 | 55.7 | 1 497 | 36 622.2 | 40.9 |
| Male |  | 236 | 6 648.0 | 35.5 | 150 | 3 158.1 | 47.5 | 236 | 5 400.0 | 43.7 | 379 | 7 028.2 | 53.9 | 1 001 | 22 234.3 | 45.0 |
| Age category |  |  |  |  |  |  |  |  |  |  |  |  |  |  |  |  |
| 34–59 |  | 162 | 9 506.3 | 17.0 | 61 | 2 348.8 | 26.0 | 106 | 4 364.2 | 24.3 | 119 | 3 568.7 | 33.3 | 448 | 19 788.0 | 22.6 |
| 60–69 |  | 238 | 6 316.0 | 37.7 | 127 | 3 312.1 | 38.3 | 178 | 4 681.7 | 38.0 | 277 | 6 044.5 | 45.8 | 820 | 20 354.3 | 40.3 |
| 70–100 |  | 213 | 4 011.8 | 53.1 | 195 | 3 126.9 | 62.4 | 292 | 4 347.1 | 67.2 | 530 | 7 228.4 | 73.3 | 1 230 | 18 714.3 | 65.7 |
| BMI category† |  |  |  |  |  |  |  |  |  |  |  |  |  |  |  |  |
| Normal |  | 503.0 | 17 183.3 | 29.3 | 280.0 | 6 930.0 | 40.4 | 410.0 | 9 778.4 | 41.9 | 613.0 | 11 247.9 | 54.5 | 1 806.0 | 45 139.7 | 40.0 |
| Overweight or obesity |  | 110.0 | 2 650.8 | 41.5 | 103.0 | 1 857.7 | 55.4 | 166.0 | 3 614.7 | 45.9 | 313.0 | 5 593.6 | 56.0 | 692.0 | 13 716.8 | 50.4 |
| Self-reported drinking status |  |  |  |  |  |  |  |  |  |  |  |  |  |  |  |  |
| Nondrinker |  | 378.5 | 12 122.9 | 31.2 | 260.9 | 5 552.7 | 47.0 | 367.1 | 8 013.9 | 45.8 | 577.6 | 9 898.6 | 58.3 | 1 584.0 | 35 588.1 | 44.5 |
| Drinker |  | 234.5 | 7 711.2 | 30.4 | 122.1 | 3 235.1 | 37.8 | 209.0 | 5 379.2 | 38.8 | 348.5 | 6 942.9 | 50.2 | 914.0 | 23 268.4 | 39.3 |
| Self-reported smoking status |  |  |  |  |  |  |  |  |  |  |  |  |  |  |  |  |
| Nonsmoker |  | 524.0 | 16 589.3 | 31.6 | 340.1 | 7 635.4 | 44.5 | 505.9 | 11 415.3 | 44.3 | 816.6 | 14 386.2 | 56.8 | 2 186.4 | 50 026.2 | 43.7 |
| Smoker |  | 89.0 | 3 244.8 | 27.4 | 43.0 | 1 152.3 | 37.3 | 70.2 | 1 977.8 | 35.5 | 109.5 | 2 455.4 | 44.6 | 311.6 | 8 830.3 | 35.3 |
| Dyslipidemia‡ |  |  |  |  |  |  |  |  |  |  |  |  |  |  |  |  |
| No |  | 290.4 | 10 423.9 | 27.9 | 172.8 | 4 165.2 | 41.5 | 225.3 | 5 659.3 | 39.8 | 374.6 | 6 697.9 | 55.9 | 1 063.1 | 26 946.2 | 39.5 |
| Yes |  | 322.6 | 9 410.2 | 34.3 | 210.3 | 4 622.6 | 45.5 | 350.7 | 7 733.8 | 45.4 | 551.4 | 10 143.7 | 54.4 | 1 434.9 | 31 910.3 | 45.0 |
| Residential district |  |  |  |  |  |  |  |  |  |  |  |  |  |  |  |  |
| East |  | 132.3 | 4 129.3 | 32.0 | 68.8 | 1 892.9 | 36.3 | 126.7 | 2 819.4 | 44.9 | 192.2 | 3 265.9 | 58.8 | 519.9 | 12 107.5 | 42.9 |
| Tatsukawa |  | 94.3 | 3 556.8 | 26.5 | 62.0 | 1 489.6 | 41.6 | 73.2 | 2 187.4 | 33.4 | 153.4 | 2 905.3 | 52.8 | 382.9 | 10 139.1 | 37.8 |
| South |  | 83.9 | 2 617.9 | 32.0 | 49.9 | 1 123.3 | 44.4 | 79.5 | 1 639.9 | 48.5 | 143.6 | 2 388.1 | 60.1 | 356.8 | 7 769.2 | 45.9 |
| Fudeoka |  | 62.8 | 2 378.7 | 26.4 | 41.5 | 1 037.9 | 40.0 | 80.7 | 1 765.5 | 45.7 | 106.5 | 2 062.7 | 51.6 | 291.5 | 7 244.9 | 40.2 |
| Central |  | 68.6 | 2 232.8 | 30.7 | 43.4 | 972.3 | 44.6 | 68.0 | 1 495.0 | 45.5 | 93.3 | 1 913.0 | 48.8 | 273.3 | 6 613.3 | 41.3 |
| Yoshiwara |  | 68.8 | 1 943.6 | 35.4 | 50.5 | 1 036.9 | 48.7 | 58.7 | 1 239.9 | 47.3 | 100.4 | 1 796.6 | 55.9 | 278.3 | 6 017.0 | 46.3 |
| West |  | 68.0 | 1 947.0 | 34.9 | 44.7 | 781.6 | 57.1 | 61.7 | 1 457.5 | 42.3 | 73.1 | 1 408.7 | 51.9 | 247.5 | 5 594.7 | 44.2 |
| Yogita |  | 34.4 | 1 028.0 | 33.4 | 22.3 | 453.4 | 49.2 | 27.8 | 788.5 | 35.2 | 63.5 | 1 101.1 | 57.7 | 148.0 | 3 371.0 | 43.9 |
| HbA1c values |  |  |  |  |  |  |  |  |  |  |  |  |  |  |  |  |
| Mean (SD) |  | 5.48 | (0.38) |  | 5.52 | (0.39) |  | 5.47 | (0.41) |  | 5.47 | (0.41) |  | 5.48 | (0.40) |  |

Abbreviations: ACC, American College of Cardiology; AHA, American Heart Association; BMI, body mass index; DBP, diastolic blood pressure; HbA1c, hemoglobin A1C; IR, incidence rate; PY, person-years; SBP, systolic blood pressure; SD, standard deviation.
Results for multiple imputed variables (overweight or obesity, self-reported drinking status, self-reported smoking status, dyslipidemia, and residential district) are averaged over 40 imputations.
\*Incidence rate is reported per 1 000 person-years.
†Overweight or obesity is defined as a BMI $\geq 25$ kg/m<sup>2</sup>.
‡Dyslipidemia is defined as serum low-density lipoprotein cholesterol $\geq 140$ mg/dL, serum high-density lipoprotein cholesterol $< 40$ mg/dL, and/or serum triglycerides $\geq 150$ mg/dL.

Table 2 shows the relationships between the ACC/AHA BP classification and the subsequent onset of CKD. In the full model, participants with elevated BP and hypertension stage 2 had a 9% (95% CI: 1%–16%) and 11% (95% CI: 5%–17%) shorter survival, respectively, than those with normal BP. Hypertension stage 1 showed shorter survival to CKD onset by point estimate but all 95% CIs crossed 1 in all models.

**Table 2.** New onset of chronic kidney disease according to the ACC/AHA blood pressure classification among 7 363 non-diabetic Japanese citizens of Zentsuji City (1998–2023).

|  |  |  |  |  |  | Crude | Model 1 | Model 2 | Model 3 | Model 4 |
| --- | --- | --- | --- | --- | --- | --- | --- | --- | --- | --- |
| Variable | SBP<br>(mmHg) |  | DBP<br>(mmHg) | PY | Failure | TR (95% CI) | aTR (95% CI) | aTR (95% CI) | aTR (95% CI) | aTR (95% CI) |
| ACC/AHA blood pressure classification |  |  |  |  |  |  |  |  |  |  |
| Normal (reference) | <120 | and | <80 | 19 834.1 | 613 | 1.00 | 1.00 | 1.00 | 1.00 | 1.00 |
| Elevated | 120–129 | and | <80 | 8 787.8 | 383 | 0.97 (0.93–1.00) | 0.91 (0.84–0.98) | 0.91 (0.84–0.98) | 0.91 (0.84–0.99) | 0.91 (0.84–0.99) |
| Hypertension stage 1 | 130–139 | or | 80–89 | 13 393.1 | 576 | 0.96 (0.93–0.99) | 0.94 (0.88–1.00) | 0.94 (0.88–1.01) | 0.95 (0.89–1.01) | 0.95 (0.89–1.01) |
| Hypertension stage 2 | ≥140 | or | ≥90 | 16 841.6 | 926 | 0.93 (0.90–0.96) | 0.87 (0.81–0.93) | 0.88 (0.83–0.95) | 0.89 (0.83–0.95) | 0.89 (0.83–0.95) |

Abbreviations: ACC, American College of Cardiology; AHA, American Heart Association; aTR: adjusted time ratio; BMI, body mass index; CI: confidence interval; DBP, diastolic blood pressure; HbA1c, glycated hemoglobin; PY, person-years; SBP, systolic blood pressure; TR, time ratio.
\*Overweight or obesity is defined as a BMI ≥25 kg/m<sup>2</sup>.
†Dyslipidemia is defined as serum low-density lipoprotein cholesterol ≥140 mg/dL, serum high-density lipoprotein cholesterol <40 mg/dL, and/or serum triglycerides ≥150 mg/dL.
Model 1: Adjusted for sex and age category (34–59[reference]/60–69/70–100). A multiplicative term (ACC/AHA blood pressure classification × age category) was added.
Model 2: Adjusted for both variables of Model 1, overweight or obesity\* (no[reference]/yes), self-reported drinking status (non- or seldom-drinker[reference]/drinker), and self-reported smoking status (non- or ex-smoker[reference]/smoker). A multiplicative term (ACC/AHA blood pressure classification × age category) was added.
Model 3: Adjusted for all variables of Model 2, dyslipidemia† (no[reference]/yes), and HbA1c values. A multiplicative term (ACC/AHA blood pressure classification × age category) was added.
Model 4: Adjuster for all variables of Model 3 and residential district (East[reference]/West/Central/South/Fudeoka/Tatsukawa/Yogita/Yoshiwara). A multiplicative term (ACC/AHA blood pressure classification × age category) was added.

### 3.2 Sensitivity analyses

In the first sensitivity analysis, we assessed the relationships between SBP and DBP levels and the development of CKD. The results using SBP categories are shown in Supplementary Table 1. In the elevated BP and hypertensive ranges, SBP of 120–129, 130–139, 140–149, and ≥150 mmHg were 19.9%, 17.9%, 12.9%, and 13.9% of the total follow-up period, respectively. Participants with SBP of 120–129 mmHg and 140–149 mmHg had an 8% (95% CI: 1%–14%) and 11% (95% CI: 3%–18%) shorter survival to the development of CKD in the fully adjusted model, respectively, than those with SBP of 100– 119 mmHg. Participants with SBP of 130–139 mmHg or SBP ≥150 mmHg had a shorter survival to CKD by point estimate, but all 95% CIs were across 1 in all models.

The relationships between DBP categories and later onset of CKD are shown in Supplementary Table 2. DBP categories of 80–89, 90–99, and ≥100 mmHg accounted for 23.6%, 8.26%, and 1.95% of the total follow-up time, respectively. There was no clear association between DBP categories and subsequent development of CKD in any of the models.

The results of the second sensitivity analysis using the ACC/AHA BP classification, which excluded participants with proteinuria ≥1+ at study entry to mitigate reverse causation, are shown in Supplementary Table 3. Compared with the follow-up times of the main analysis shown in Table 1, the follow-up times of the second sensitivity analysis were reduced by 2.33%, 3.15%, 3.06%, and 4.14% in the order of the ACC/AHA BP classification of normal, elevated, hypertension stage 1, and hypertension stage 2, respectively. The total follow-up time was the shortest in the elevated BP category (14.9% of the total follow-up time). The second sensitivity analysis yielded similar results to those of the main analysis shown in Table 2. Therefore, no clear evidence of reverse causation was confirmed.

## 4 Discussion

This longitudinal study of 7 363 non-diabetic, middle-aged and older Japanese adults (men: 40.3%) living in Zentsuji City with a mean follow-up of 7.99 years showed the following. An elevated BP classification and hypertension stage 2 classification resulted in a shorter survival to the onset of CKD than a normal BP classification (Table 2).

A previous study followed 10 096 non-diabetic, middle-aged American participants in the Atherosclerosis Risk in Communities study for 9 years[13]. In this previous study, hypertension, which was defined as SBP/DBP ≥130/≥85 mmHg or BP medication use, was associated with a 1.99-fold risk (95% CI: 1.69–2.35) of developing CKD compared with normal BP (SBP/DBP <130/<85 mmHg and no use of BP medication). In our study using the ACC/AHA classification, abnormal BP was categorized into three categories of elevated BP, hypertension stage 1, and hypertension stage 2[15]. We found that all abnormal BP categories had a shorter survival to CKD onset by point estimate, but all 95% CIs for hypertension stage 1 crossed 1 in all models (Table 2). When we used SBP (Supplementary Table 1) and DBP (Supplementary Table 2) as exposure variables in the first sensitivity analysis, only SBP of 120–129 and 140–149 mmHg were associated with new-onset CKD, similar to the main analysis in Table 2.

This may be partly explained by the statistical model we used and the unobserved information on using BP-lowering medications. First, we selected the time-varying model because the PH assumption was violated. In a previous cohort study of 7,343 non-diabetic and diabetic Korean adults without using BP-lowering medication at baseline, the hazard ratio (HR) of developing CKD was higher in all elevated SBP categories (120–129, 130–139, 140–159, ≥160 mmHg) when using the PH model, after adjustment for diabetes. However, using the time-varying model, point estimates were lower than those of the PH model estimates for all elevated SBP categories[38]. This partly explains why our results show higher point estimates (i.e., longer survival) than previous studies[5].

Second, information on the use of BP-lowering medications was not available in this study, which may have influenced the results. In a cohort study of 154 692 non-diabetic and diabetic Japanese adults, compared to normotensives not taking BP-lowering medications, hypertension stages 1 and 2 taking BP-lowering medications had higher HRs for developing CKD than hypertension stages 1 and 2 not taking BP-lowering medications, after controlling diabetes[39]. This study includes participants using and not using BP-lowering medications. According to the 8^th^ National Database in Japan, 21.2% (24.2% of men and 17.4% of women) of those aged 40–74 years (15 786 772 men and 13 009 475 women) who underwent checkups in the fiscal year 2020 self-reported taking BP-lowering medications[18,40]. This low use of BP-lowering medications, especially among women, suggests that the results of this study, in which the proportion of women was 59.7%, are underestimated and that survival to CKD onset may be shorter in all ranges of abnormal BP (Figure 1).

When we evaluated the relationship between the ACC/AHA BP classification and the onset of CKD after minimizing reverse causation in the second sensitivity analysis (excluding participants with proteinuria ≥1+ at study entry), there was no clear evidence of reverse causation. The reason for this finding is that the results were similar to those of the main analysis shown in Table 2 (Supplementary Table 3). In the non-diabetic Asian adult population, elevated BP, hypertension stage 1, and hypertension stage 2 had a high risk of a shorter time to the onset of CKD by point estimate, with 95% CIs across 1 for hypertension stage 1. This implies that it may be appropriate to consider a lower cutoff value than the current hypertension criteria (SBP ≥130 or DBP ≥80 mmHg) as a risk factor for the new onset of CKD in healthy Asian adults without diabetes.

### 4.1 Strengths and Limitations

One of the strengths of our study is its detailed examination of the relationship between BP levels and new onset of CKD in a non-diabetic, middle-aged and older Asian population using data for approximately 25 years. The time frame of our data may be long enough to detect the early stage of CKD.

However, this study has several limitations. First, the results cannot be generalized because the data were health checkup data from a single city in Japan[16]. Second, there may have been selection bias because our participants decided by themselves whether to undergo a checkup, and the following variables, which may have caused competing risks, were unavailable in this study: death, hospitalization, and the prevalence of other diseases. Our participants are likely to be healthier than the general population because they did not tend to have characteristics, such as having a serious illness that prevented them from going out, did not feel the need for further checkups because they already had a disease and regularly visited a physician, and did not consider a checkup important because they were not interested in improving their health. This situation may have caused an underestimation of the results.

Third, there was measurement error because we estimated renal function using the revised Japanese equation[20–22]. Furthermore, inulin clearance values, which are considered the gold standard for measuring the GFR, were unavailable in this study owing to the time and cost involved and the nature of the annual health checkup conducted by the local government. Fourth, unobserved variables (a family history of CKD, a family history of heart disease, and socioeconomic factors) and variables with too many missing values to include in the model (antihypertensive medication use, a history of heart disease, and a history of stroke) may have influenced the present results. Fifth, the unavailability of mortality data related to hypertension, such as cardiovascular disease, heart failure, and stroke, may have affected the inference of causality related to hypertension outcomes.

Sixth, there may have been reverse causation. The second sensitivity analysis attempted to minimize reverse causation, but did not provide evidence that it existed (Supplementary Table 2). Finally, we cannot rule out the possibility of having a built-in selection bias in which less susceptible participants remained in the cohort because of the long follow-up of approximately 25 years[41]. Therefore, all results of this study may have been underestimated.

## 5 Conclusion

In conclusion, we investigated the relationship between the ACC/AHA BP classification and the development of CKD in a middle-aged and older Japanese population without diabetes. This study showed that point estimates of survival to the development of CKD onset were shorter for elevated BP, hypertension stage 1, and hypertension stage 2 than normal BP. These findings suggest that controlling BP in the appropriate range reduces the risk of developing CKD in a relatively healthier non-diabetic population than the general population.

## Data Availability

All data produced in the present work are contained in the manuscript and its supplementary information.

## 6 Acknowledgments

The authors are grateful to all participants of this study, Ayaka Nakatsu, Masako Matsumoto, Mayumi Kitadani, and all local government officers of Zentsuji City for their support and contribution. We thank Ellen Knapp, PhD, from Edanz (https://jp.edanz.com/ac) for editing a draft of this manuscript.

## 7 Author contributions

**YO:** Conceptualization; Data curation; Formal analysis; Investigation; Methodology; Project administration; Visualization; Roles/Writing - original draft; and Writing - review & editing. **TM**: Methodology; Supervision; and Writing - review & editing. Both authors read and approved the final manuscript.

## Notes

### Competing Interest Statement

Employment: YO (Zentsuji City)

### Author Declarations

All data were retrieved from the administrative database of Zentsuji City and anonymized prior to receipt. The Ethics Committee of Okayama University Graduate School of Medicine, Dentistry and Pharmaceutical Sciences and Okayama University Hospital approved this study and waived the need for informed consent (No. K1708-040).

### Summary of Updates

The Methods and Discussion sections were revised following the reviewers' comments.

## References

[1] Bikbov B, Purcell CA, Levey AS, Smith M, Abdoli A, Abebe M, et al. Global, regional, and national burden of chronic kidney disease, 1990–2017: a systematic analysis for the Global Burden of Disease Study 2017. The Lancet 2020;395:709–33. 10.1016/S0140-6736(20)30045-3.

[2] Stevens PE, Ahmed SB, Carrero JJ, Foster B, Francis A, Hall RK, et al. KDIGO 2024 Clinical practice guideline for the evaluation and management of chronic kidney disease. Kidney International 2024;105:S117–314. 10.1016/j.kint.2023.10.018.

[3] Liyanage T, Ninomiya T, Jha V, Neal B, Patrice HM, Okpechi I, et al. Worldwide access to treatment for end-stage kidney disease: a systematic review. The Lancet 2015;385:1975–82. 10.1016/S0140-6736(14)61601-9.

[4] Shen Y, Cai R, Sun J, Dong X, Huang R, Tian S, et al. Diabetes mellitus as a risk factor for incident chronic kidney disease and end-stage renal disease in women compared with men: A systematic review and meta-analysis. Endocrine 2017;55:66–76. 10.1007/s12020-016-1014-6.

[5] Weldegiorgis M, Woodward M. The impact of hypertension on chronic kidney disease and end-stage renal disease is greater in men than women: a systematic review and meta-analysis. BMC Nephrol 2020;21:506. 10.1186/s12882-020-02151-7.

[6] Fabrizi F, Cerutti R, Donato FM, Messa P. HBV infection is a risk factor for chronic kidney disease: Systematic review and meta-analysis. Revista Clínica Española (English Edition) 2021;221:600–11. 10.1016/j.rceng.2019.10.014.

[7] Fabrizi F, Verdesca S, Messa P, Martin P. Hepatitis C virus infection increases the risk of developing chronic kidney disease: a systematic review and meta-analysis. Dig Dis Sci 2015;60:3801–13. 10.1007/s10620-015-3801-y.

[8] He L-Q, Wu X-H, Huang Y-Q, Zhang X-Y, Shu L. Dietary patterns and chronic kidney disease risk: a systematic review and updated meta-analysis of observational studies. Nutr J 2021;20:4. 10.1186/s12937-020-00661-6.

[9] Okoye OC, Carnegie E, Mora L. Air pollution and chronic kidney disease risk in oil and gas- situated communities: a systematic review and meta-analysis. Int J Public Health 2022;67:1604522. 10.3389/ijph.2022.1604522.

[10] Cao X, Xie X, Zhou J, Yuan H, Chen Z. Relationship between prehypertension and incidence of chronic kidney disease in a general population: a prospective analysis in central south China. Int Urol Nephrol 2014;46:2183–9. 10.1007/s11255-014-0805-z.

[11] Jee SH, Boulware LE, Guallar E, Suh I, Appel LJ, Miller ER. Direct, progressive association of cardiovascular risk factors with incident proteinuria: results from the Korea Medical Insurance Corporation (KMIC) Study. Arch Intern Med 2005;165:2299. 10.1001/archinte.165.19.2299.

[12] Tohidi M, Hasheminia M, Mohebi R, Khalili D, Hosseinpanah F, Yazdani B, et al. Incidence of chronic kidney disease and its risk factors, results of over 10 year follow up in an Iranian cohort. PLoS ONE 2012;7:e45304. 10.1371/journal.pone.0045304.

[13] Kurella M, Lo JC, Chertow GM. Metabolic syndrome and the risk for chronic kidney disease among nondiabetic adults. Journal of the American Society of Nephrology 2005;16:2134–40. 10.1681/ASN.2005010106.

[14] Aggarwal R, Chiu N, Wadhera RK, Moran AE, Raber I, Shen C, et al. Racial/ethnic disparities in hypertension prevalence, awareness, treatment, and control in the United States, 2013 to 2018. Hypertension 2021;78:1719–26. 10.1161/HYPERTENSIONAHA.121.17570.

[15] Whelton PK, Carey RM, Aronow WS, Casey DE, Collins KJ, Dennison Himmelfarb C, et al. 2017 ACC/AHA/AAPA/ABC/ACPM/AGS/APhA/ASH/ASPC/NMA/PCNA guideline for the prevention, detection, evaluation, and management of high blood pressure in adults: Executive summary: a report of the American college of cardiology/American heart association task force on clinical practice guidelines. Hypertension 2018;71:1269–324. 10.1161/HYP.0000000000000066.

[16] Zentsuji City. Zentsuji City official website n.d. https://www.city.zentsuji.kagawa.jp (accessed April 18, 2024).

[17] Okawa Y, Suzuki E, Mitsuhashi T, Tsuda T, Yorifuji T. A population-based longitudinal study on glycated hemoglobin levels and new-onset chronic kidney disease among non-diabetic Japanese adults. Sci Rep 2023;13:13770. 10.1038/s41598-023-40300-8.

[18] Ministry of Health, Labour and Welfare. Specific health examination and specific health guidance n.d. https://www.mhlw.go.jp/stf/seisakunitsuite/bunya/0000161103.html (accessed April 18, 2024).

[19] ElSayed NA, Aleppo G, Aroda VR, Bannuru RR, Brown FM, Bruemmer D, et al. 2. Classification and diagnosis of diabetes: *standards of care in diabetes—*2023. Diabetes Care 2023;46:S19–40. 10.2337/dc23-S002.

[20] Matsuo S, Imai E, Horio M, Yasuda Y, Tomita K, Nitta K, et al. Revised equations for estimated GFR from serum creatinine in Japan. American Journal of Kidney Diseases 2009;53:982–92. 10.1053/j.ajkd.2008.12.034.

[21] Matsuo S, Yasuda Y, Imai E, Horio M. Current status of estimated glomerular filtration rate (eGFR) equations for Asians and an approach to create a common eGFR equation: approach to eGFR equation for Asians. Nephrology 2010;15:45–8. 10.1111/j.1440-1797.2010.01313.x.

[22] Fujii R, Pattaro C, Tsuboi Y, Ishihara Y, Melotti R, Yamada H, et al. Comparison of glomerular filtration rate estimating formulas among Japanese adults without kidney disease. Clinical Biochemistry 2023;111:54–9. 10.1016/j.clinbiochem.2022.10.011.

[23] World Health Organization. A healthy lifestyle - WHO recommendations n.d. https://www.who.int/europe/news-room/fact-sheets/item/a-healthy-lifestyle---who-recommendations (accessed April 18, 2024).

[24] Kinoshita M, Yokote K, Arai H, Iida M, Ishigaki Y, Ishibashi S, et al. Japan Atherosclerosis Society (JAS) guidelines for prevention of atherosclerotic cardiovascular diseases 2017. J Atheroscler Thromb 2018;25:846–984. 10.5551/jat.GL2017.

[25] NGSP. NGSP Home n.d. https://ngsp.org (accessed April 18, 2024).

[26] Kashiwagi A, Kasuga M, Araki E, Oka Y, Hanafusa T, Ito H, et al. International clinical harmonization of glycated hemoglobin in Japan: from Japan Diabetes Society to National Glycohemoglobin Standardization Program values. Journal of Diabetes Investigation 2012;3:39–40. 10.1111/j.2040-1124.2012.00207.x.

[27] Young R, Johnson DR. Handling missing values in longitudinal panel data with multiple imputation. Fam Relat 2015;77:277–94. 10.1111/jomf.12144.

[28] von Hippel PT. How many imputations do you need? A two-stage calculation using a quadratic rule. Sociological Methods & Research 2020;49:699–718. 10.1177/0049124117747303.

[29] Schoenfeld D. Partial residuals for the proportional hazards regression model. Biometrika 1982;69:239–41. 10.1093/biomet/69.1.239.

[30] Akaike H. A new look at the statistical model identification. IEEE Trans Automat Contr 1974;19:716–23. 10.1109/TAC.1974.1100705.

[31] Schwarz G. Estimating the dimension of a model. Ann Statist 1978;6. 10.1214/aos/1176344136.

[32] Westreich D, Greenland S. The Table 2 fallacy: presenting and interpreting confounder and modifier coefficients. American Journal of Epidemiology 2013;177:292–8. 10.1093/aje/kws412.

[33] National Statistics Center of Japan. Portal Site of Official Statistics of Japan. Portal Site of Official Statistics of Japan n.d. https://www.e-stat.go.jp/en (accessed April 18, 2024).

[34] Delker CJ. Schemdraw 0.18 documentation n.d. https://schemdraw.readthedocs.io/en/latest/ (accessed January 30, 2024).

[35] Bossche JV den, Fleischmann M, McBride J, Ward B, Wolf L, Richards M. GeoPandas 0.14.2 documentation n.d. https://geopandas.org/en/stable/index.html (accessed April 18, 2024).

[36] Python Software Foundation. Python. PythonOrg 2023. https://www.python.org/ (accessed April 18, 2024).

[37] Vandenbroucke JP, von Elm E, Altman DG, Gøtzsche PC, Mulrow CD, Pocock SJ, et al. Strengthening the reporting of observational studies in epidemiology (STROBE): explanation and elaboration. Epidemiology 2007;18:805–35. 10.1097/EDE.0b013e3181577511.

[38] Lee H, Kwon SH, Jeon JS, Noh H, Han DC, Kim H. Association between blood pressure and the risk of chronic kidney disease in treatment-naïve hypertensive patients. Kidney Res Clin Pract 2022;41:31–42. 10.23876/j.krcp.21.099.

[39] Satoh M, Hirose T, Nakayama S, Murakami T, Takabatake K, Asayama K, et al. Blood pressure and chronic kidney disease stratified by gender and the use of antihypertensive drugs. JAHA 2020;9:e015592. 10.1161/JAHA.119.015592.

[40] Ministry of Health, Labour and Welfare. NDB Open Data. NDB Open Data n.d. https://www.mhlw.go.jp/stf/seisakunitsuite/bunya/0000177182.html (accessed April 18, 2024).

[41] Hernán MA. The hazards of hazard ratios. Epidemiology 2010;21:13–5. 10.1097/EDE.0b013e3181c1ea43.

