## Supplementary Tables for "Association between blood pressure and new onset of chronic kidney disease in non-diabetic Japanese adults: a population-based longitudinal study from 1998 to 2023"

| **Supplementary Table 1.** New onset of chronic kidney disease according to the SBP category among 7 363 non-diabetic Japanese citizens of Zentsuji City (1998–2023). | | | | | | | |
| --- | --- | --- | --- | --- | --- | --- | --- |
|  |  |  | Crude | Model 1 | Model 2 | Model 3 | Model 4 |
| Variable | PY | Failure | TR (95% CI) | aTR (95% CI) | aTR (95% CI) | aTR (95% CI) | aTR (95% CI) |
| SBP category | | | | | | | |
| <100 mmHg | 3 825.7 | 78 | 1.10 (1.03–1.18) | 1.16 (1.03–1.30) | 1.15 (1.02–1.29) | 1.15 (1.02–1.29) | 1.15 (1.02–1.29) |
| 100–119 mmHg (reference) | 17 009.2 | 565 | 1.00 | 1.00 | 1.00 | 1.00 | 1.00 |
| 120–129 mmHg | 11 714.6 | 498 | 0.98 (0.94–1.01) | 0.92 (0.86–0.98) | 0.92 (0.86–0.99) | 0.92 (0.86–0.99) | 0.92 (0.86–0.99) |
| 130–139 mmHg | 10 525.5 | 478 | 0.97 (0.94–1.01) | 0.96 (0.89–1.04) | 0.97 (0.90–1.04) | 0.97 (0.90–1.05) | 0.97 (0.90–1.05) |
| 140–149 mmHg | 7 593.8 | 403 | 0.95 (0.91–0.98) | 0.88 (0.81–0.96) | 0.89 (0.82–0.97) | 0.89 (0.82–0.97) | 0.89 (0.82–0.97) |
| ≥150 mmHg | 8 187.7 | 476 | 0.94 (0.90–0.97) | 0.91 (0.83–1.00) | 0.92 (0.84–1.01) | 0.92 (0.84–1.01) | 0.92 (0.84–1.01) |
| Abbreviations: aTRs: adjusted time ratios; CI: confidence interval; BMI, body mass index; HbA1c, glycated hemoglobin; PY, person-years; SBP, systolic blood pressure; TR, time ratio. | | | | | | | |
| *Overweight or obesity is defined as a BMI ≥25 kg/m^2^. | | | | | | | |
| †Dyslipidemia is defined as serum low-density lipoprotein cholesterol ≥140 mg/dL, serum high-density lipoprotein cholesterol <40 mg/dL, and/or serum triglycerides ≥150 mg/dL. | | | | | | | |
| Model 1: Adjusted for sex and age category (34–59[reference]/60–69/70–100). A multiplicative term (SBP category × age category) was added. | | | | | | | |
| Model 2: Adjusted for both variables of Model 1, overweight or obesity* (no[reference]/yes), self-reported drinking status (non- or seldom-drinker[reference]/drinker), and self-reported smoking status (non- or ex-smoker[reference]/smoker). A multiplicative term (SBP category × age category) was added. | | | | | | | |
| Model 3: Adjusted for all variables of Model 2, dyslipidemia† (no[reference]/yes), and HbA1c values. A multiplicative term (SBP category × age category) was added. | | | | | | | |
| Model 4: Adjuster for all variables of Model 3 and residential district (East[reference]/West/Central/South/Fudeoka/Tatsukawa/Yogita/Yoshiwara). A multiplicative term (SBP category × age category) was added. | | | | | | | |

| **Supplementary Table 2.** New onset of chronic kidney disease according to the DBP category among 7 363 non-diabetic Japanese citizens of Zentsuji City (1998–2023). | | | | | | | |
| --- | --- | --- | --- | --- | --- | --- | --- |
|  |  |  | Crude | Model 1 | Model 2 | Model 3 | Model 4 |
| Variable | PY | Failure | TR (95% CI) | aTR (95% CI) | aTR (95% CI) | aTR (95% CI) | aTR (95% CI) |
| DBP category | | | | | | | |
| <70 mmHg | 18 982.0 | 680 | 1.04 (1.01–1.06) | 1.12 (1.05–1.20) | 1.11 (1.04–1.19) | 1.11 (1.03–1.18) | 1.11 (1.03–1.18) |
| 70–79 mmHg (reference) | 19 971.2 | 882 | 1.00 | 1.00 | 1.00 | 1.00 | 1.00 |
| 80–89 mmHg | 13 890.2 | 633 | 0.99 (0.96–1.01) | 0.99 (0.93–1.06) | 0.98 (0.91–1.04) | 0.98 (0.91–1.04) | 0.98 (0.91–1.04) |
| 90–99 mmHg | 4 863.6 | 247 | 0.95 (0.92–0.99) | 0.96 (0.88–1.04) | 0.97 (0.88–1.05) | 0.97 (0.89–1.06) | 0.97 (0.89–1.06) |
| ≥100 mmHg | 1 149.5 | 56 | 0.94 (0.88–1.01) | 0.98 (0.85–1.12) | 0.99 (0.85–1.14) | 0.99 (0.85–1.14) | 0.99 (0.85–1.15) |
| Abbreviations: aTRs: adjusted time ratios; CI: confidence interval; BMI, body mass index; DBP, diastolic blood pressure; HbA1c, glycated hemoglobin; PY, person-years; TR, time ratio. | | | | | | | |
| *Overweight or obesity is defined as a BMI ≥25 kg/m^2^. | | | | | | | |
| †Dyslipidemia is defined as serum low-density lipoprotein cholesterol ≥140 mg/dL, serum high-density lipoprotein cholesterol <40 mg/dL, and/or serum triglycerides ≥150 mg/dL. | | | | | | | |
| Model 1: Adjusted for sex and age category (34–59[reference]/60–69/70–100). A multiplicative term (DBP category × age category) was added. | | | | | | | |
| Model 2: Adjusted for both variables of Model 1, overweight or obesity* (no[reference]/yes), self-reported drinking status (non- or seldom-drinker[reference]/drinker), and self-reported smoking status (non- or ex-smoker[reference]/smoker). The multiplicative terms (DBP category × age category, DBP × overweight or obese*) were added. | | | | | | | |
| Model 3: Adjusted for all variables of Model 2, dyslipidemia† (no[reference]/yes), and HbA1c values. The multiplicative terms (DBP category × age category, DBP × overweight or obese*) were added. | | | | | | | |
| Model 4: Adjuster for all variables of Model 3 and residential district (East[reference]/West/Central/South/Fudeoka/Tatsukawa/Yogita/Yoshiwara). The multiplicative terms (DBP category × age category, DBP × overweight or obese*) were added. | | | | | | | |

| **Supplementary Table 3.** New onset of chronic kidney disease according to the ACC/AHA blood pressure classification among 7 077 non-diabetic Japanese citizens of Zentsuji City excluding those with proteinuria ≥1+ at study entry (1998–2023). | | | | | | | | | | |
| --- | --- | --- | --- | --- | --- | --- | --- | --- | --- | --- |
|  |  |  |  |  |  | Crude | Model 1 | Model 2 | Model 3 | Model 4 |
| Variable | SBP (mmHg) |  | DBP (mmHg) | PY | Failure | TR (95% CI) | aTR (95% CI) | aTR (95% CI) | aTR (95% CI) | aTR (95% CI) |
| ACC/AHA blood pressure classification | | | | | | | | | | |
| Normal (reference) | <120 | and | <80 | 19 383.2 | 597 | 1.00 | 1.00 | 1.00 | 1.00 | 1.00 |
| Elevated | 120–129 | and | <80 | 8 519.7 | 364 | 0.97 (0.94–1.01) | 0.91 (0.84–0.99) | 0.92 (0.85–1.00) | 0.92 (0.84–0.99) | 0.92 (0.84–0.99) |
| Hypertension stage 1 | 130–139 | or | 80–89 | 12 995.2 | 551 | 0.96 (0.93–0.99) | 0.94 (0.88–1.00) | 0.93 (0.87–1.00) | 0.95 (0.89–1.01) | 0.95 (0.89–1.01) |
| Hypertension stage 2 | ≥140 | or | ≥90 | 16 171.5 | 878 | 0.93 (0.90–0.96) | 0.87 (0.81–0.94) | 0.87 (0.81–0.93) | 0.89 (0.83–0.95) | 0.89 (0.83–0.95) |
| Abbreviations: ACC, American College of Cardiology; AHA, American Heart Association; aTR: adjusted time ratio; BMI, body mass index; CI: confidence interval; DBP, diastolic blood pressure; HbA1c, glycated hemoglobin; PY, person-years; SBP, systolic blood pressure; TR, time ratio. | | | | | | | | | | |
| *Overweight or obesity is defined as a BMI ≥25 kg/m^2^. | | | | | | | | | | |
| †Dyslipidemia is defined as serum low-density lipoprotein cholesterol ≥140 mg/dL, serum high-density lipoprotein cholesterol <40 mg/dL, and/or serum triglycerides ≥150 mg/dL. | | | | | | | | | | |
| Model 1: Adjusted for sex and age category (34–59[reference]/60–69/70–100). A multiplicative term (ACC/AHA blood pressure classification × age category) was added. | | | | | | | | | | |
| Model 2: Adjusted for both variables of Model 1, overweight or obesity* (no[reference]/yes), self-reported drinking status (non- or seldom-drinker[reference]/drinker), and self-reported smoking status (non- or ex-smoker[reference]/smoker). The multiplicative terms (ACC/AHA blood pressure classification × age category, ACC/AHA blood pressure classification × overweight or obesity*) were added. | | | | | | | | | | |
| Model 3: Adjusted for all variables of Model 2, dyslipidemia† (no[reference]/yes), and HbA1c values. A multiplicative term (ACC/AHA blood pressure classification × age category) was added. | | | | | | | | | | |
| Model 4: Adjuster for all variables of Model 3 and residential district (East[reference]/West/Central/South/Fudeoka/Tatsukawa/Yogita/Yoshiwara). A multiplicative term (ACC/AHA blood pressure classification × age category) was added. | | | | | | | | | | |
